# Simplified Drainless Outpatient Female-to-Male Gender-Affirming Bilateral Mastectomy

**DOI:** 10.1101/2022.06.19.22271559

**Authors:** Sean A. Knudson, Ashley DeLeon, Curtis N. Crane, Richard A. Santucci

**Affiliations:** Crane Center for Transgender Surgery; Austin, TX

**Author notes:** **Corresponding Author**: Richard A. Santucci, MD, HON FC Urol(SA), FACS, Crane Center for Transgender Surgery, 5656 Bee Cave Rd Suite J201, Austin, TX 78746. Financial Disclosure Statement: Mr. Knudson, Dr. DeLeon, Dr. Crane, and Dr. Santucci have nothing to disclose. No funding was received for this article. None of the authors has a financial interest in any of the products, devices, or drugs mentioned in this manuscript. Institutional Review Board Approval/Declaration of Helsinki Statement: This study has was conducted with IRB approval and all authors adhered to the Declaration of Helsinki throughout the entirety of the study period.

## Abstract

**Purpose:** Female-to-male gender-affirming top surgery is growing in demand. We ventured to further improve double-incision free nipple graft bilateral mastectomy by utilizing a streamlined method of eliminating dead space and abandoning the practice of postoperative drain placement.

**Methods:** Patients with gender dysphoria and who underwent streamlined gender-affirming top surgery without drain placement were retrospectively reviewed from August 2017 to June 2020. A literature review was conducted to identify comparative studies with historical complication data. Patient outcomes were analyzed against this aggregated data.

**Results:** One-hundred and seven patients underwent 214 simplified double incision free nipple graft bilateral mastectomies in an outpatient surgery center. Mean patient age was 27.2 ± 10.4 years. The overall complication rate was 13.1 percent. Hematoma occurred in 2 patients (1.9%). Seroma occurred in 10 patients (9.3%). Wound dehiscence occurred in 2 patients (1.9%). Elective revision rate was 3/107 (2.8%). One patient had acute reoperation due to major hematoma (0.9%). Compared with eleven studies of pooled historical outcomes of patients with drain placement, analysis revealed the drainless group had significantly higher rates of seroma (*p* = 0.003353), but significantly lower rates of revision (*p* = 1.37×10^−12^). Aggregation of our data with two past drainless studies was compared to the eleven drain inclusive studies, revealing significantly lower rates of hematoma (*p* = 0.001069), nipple areola complex necrosis (*p* = 0.01034), and revision (*p* = 2.20×10^−16^).

**Conclusion:** Simplified, drainless, outpatient double incision free nipple graft bilateral mastectomy can be performed with comparable outcomes to historical data.

## Introduction

Female-to-male (FTM) gender-affirming bilateral mastectomy (“top surgery”) is a significant and common step for individuals with gender dysphoria transitioning to a masculine phenotype. Surgical facilitation of physical alignment with one’s gender identity can have a notable and long-term psychological and aesthetic impact by benefiting patient well-being, sexual function, and bodily acceptance.^1–4^ Globally, there is an estimated 0.3 to 0.5 percent prevalence of transgenderism,^5^ and it has been reported that 1.4 million adults (0.6 percent) in the United States identify as transgender as of 2016.^6^ Clinical demand for, and insurance coverage of, gender-affirming surgery has substantially increased in the last half-decade,^7^ further increasing demand for safe and effective procedures. Despite a numerical increase in procedures, there still is a deficit in best practice and outcome literature in this field.

Since the first surgical case series published in the 1990’s,^8^ there has been increased attention to improved top surgery techniques. Initially, it was standard for surgeons to include drain placement after both standard mastectomy (for cancer), and double-incision free nipple graft (DIFNG) bilateral mastectomy (for gender affirmation surgery).^9,10,19,11–18^ In the early 2000’s, authors began reporting success after drainless radical mastectomy performed for cancer, suggesting that surgical drains were not required for a good result.^20^ Recently in a series of 153 patients, Gallagher et al. reported good results with a drainless bilateral mastectomy technique for gender affirmation when “as much dead space as possible [was] obliterated using a progressive tension technique similar to that described by Pollock and Pollock in abdominoplasty”.^21^

We endeavored to further improve and simplify drainless DIFNG top surgery by streamlining the method of dead space obliteration, confirming the observations of Gallagher^21^ and McEvenue^22^ that drains need not be used in gender affirming mastectomy, and establishing that this surgery can be safely performed on an outpatient basis. We hypothesized that our simplified, drainless DIFNG top surgery technique would yield similar results as surgery that utilizes drains.

### Patients & Methods

We performed a retrospective cohort review of patients who underwent bilateral DIFNG mastectomy for masculinizing top surgery between August 2017 and June 2020. The electronic medical records of all patients with diagnosis of gender dysphoria and who underwent DIFNG were reviewed. Patients that underwent circumareolar incision or “keyhole” technique were excluded. Relevant epidemiologic data and clinical details were collected, including: age, body mass index (BMI), obesity, race, smoking history, testosterone usage, and medical history. Complications recorded were: seroma, hematoma, dehiscence, surgical site infection (SSI), nipple areola complex (NAC) necrosis, and revision surgery. Operative data included: estimated blood loss and resection mass of the right and left breasts. Major complications were defined as any adverse condition that required a return to the operating room. Minor complications were those that did not require a return to the operating room, and were either monitored during routine follow-up in the office or resolved on their own. Patients were followed up 1-2 weeks after surgery. Healthcare data was recorded including: policy type, healthcare provider, and patient co-pay.

#### Comparative Studies

A literature review of PubMed® was conducted to select for comparative outcome studies that described DIFNG with drain placement and that described DIFNG without drain placement. Search terms included: “male-to-female,” “mastectomy,” “top surgery,” and “chest contouring.” Studies that were not retrospective reviews of analyses of FTM top surgery were not included. If studies used multiple mastectomy techniques, only outcomes of DIFNG technique were used for our analysis. Outcomes in our study were compared to the historical data of studies where drains were used and were not used.

#### Data Analysis

Analysis of the sample size included tabulating the above variables using the descriptive statistics of mean, standard deviation (SD), minimum, mode, frequency, and percentages. Continuous variables were expressed using “mean ± SD (range),” and categorical variables were expressed using frequencies and percentages. Statistics used to describe categorical variables were compared with congruous statistics in the current plastic surgery outcomes literature using Fisher’s exact test in RStudio Version 1.3.1093 (RStudio, PBC, Boston, MA). Significance was set at *p* < 0.05.

#### Streamlined and Simplified Double Incision Free Nipple Graft Technique

Preoperatively, the inframammary fold, at a point slightly below the pectoralis muscle, is marked in the sitting position with the breast lifted. The breast is then placed in the neutral resting position and the inframammary marking is transposed to the ventral surface of the breast (Figure 1). This ellipse of skin created by the inferior chest mark and the more superior mark on the underside of the breast is later removed during mastectomy.

**Figure 1.**
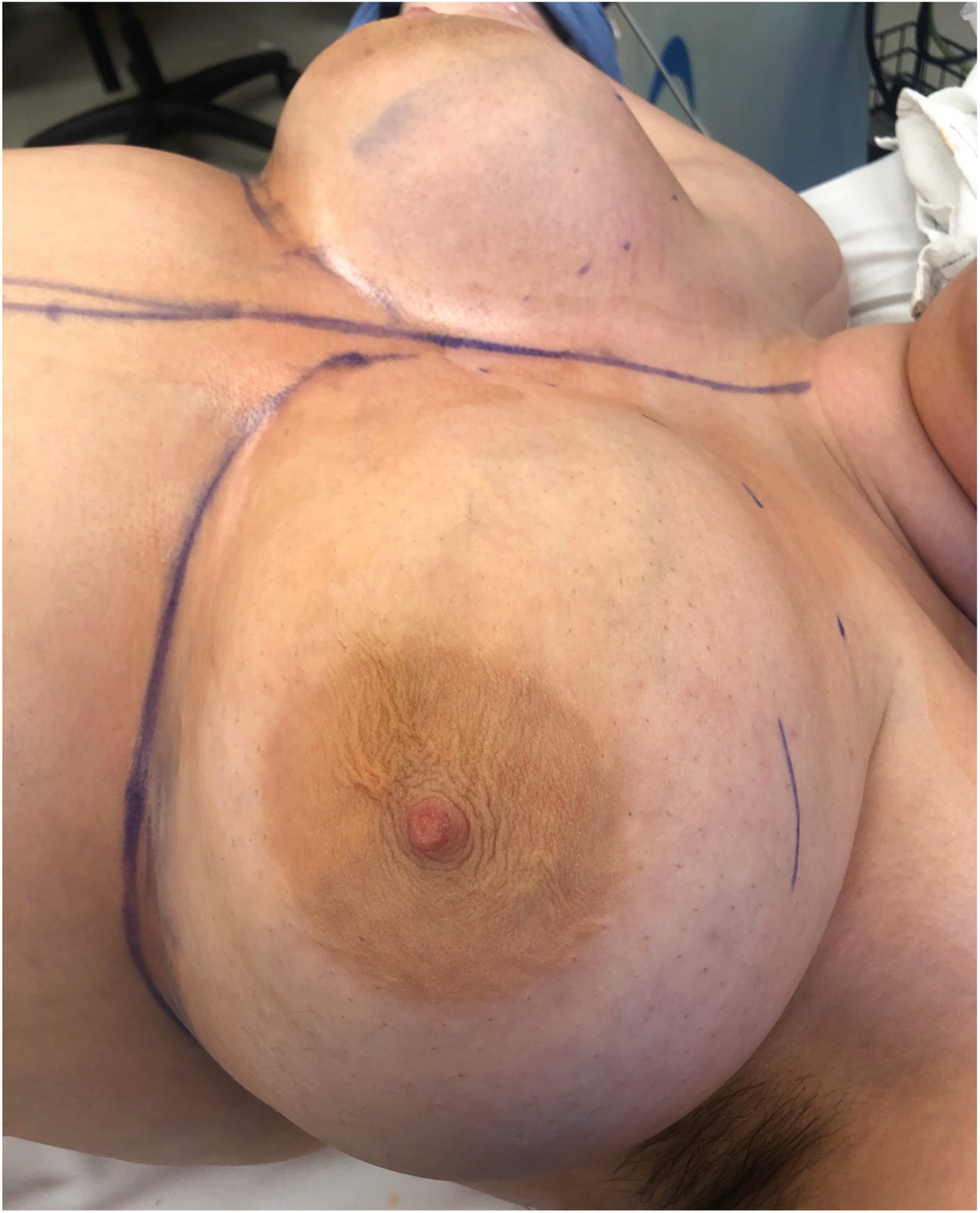
Preoperative photo showing marked breast with upper incision line noted on the upper breast.

First the nipples are removed and thinned. Bilateral inframammary incisions are made slightly below the lateral margin of the pectoralis major and carried down to the chest wall with electrocautery (40W) – following the inferior contour of the pectoralis major rather than the inframammary fold. The incision is carried cephalad along the pectoralis fascia, and extended superiorly and laterally to separate the breast tissue from the fascia. The skin is pulled taught, insuring that the elliptical incision will close with minimal skin tension. The ventrally marked incision is made. The superior aspect of the mammary tissue is dissected away from the subcutaneous fat at the breast capsule, following cephalad until the breast tissue is completely removed.

The superficial fascia of the superior flap is then closed to pectoralis fascia with two 0-0 Vicryl (polyglactin) simple interrupted sutures, one at midflap and one at the anterior axillary line, for a total of 4 sutures bilaterally (Figure 2, Figure 3). The Insorb® absorbable stapler system (Cooper Surgical; Trumball, CT, USA) is used to place 15 absorbable staples on each side. The device delivers 30 smooth, non-porous absorbable staples comprised of a co-polymer of predominantly polylactic acid, and is equipped with a proprietary delivery device that draws the tissue together before delivering the staple. To avoid medial or lateral dog ear (which can be especially easy to create because the superior and inferior incisions are of different lengths) the staples are placed medially, then laterally, then in the middle to achieve equal closure throughout the incision. Roughly half of the staples are placed deep to dermis and serve to further obliterate dead space without puckering the skin (Figure 4). After roughly half the internal staples are placed, 2 additional 0-0 Vicryl sutures are placed in the nearly-closed wound to further affix the superficial fascia of the flap to the pectoralis fascia, again being careful not to pucker the skin. The remaining absorbable staples are placed into the wound to complete deep closure. The nipple-areolar complexes are harvested as single-unit full-thickness skin grafts and affixed using standard, described techniques and bolster dressings.^22^ The skin is closed with subcuticular 3-0 Monocryl (poliglecaprone 25) (Figure 5), dressed with Mastisol liquid adhesive and Steri-Stips, and the chest is wrapped with an abdominal binder.

**Figure 2.**
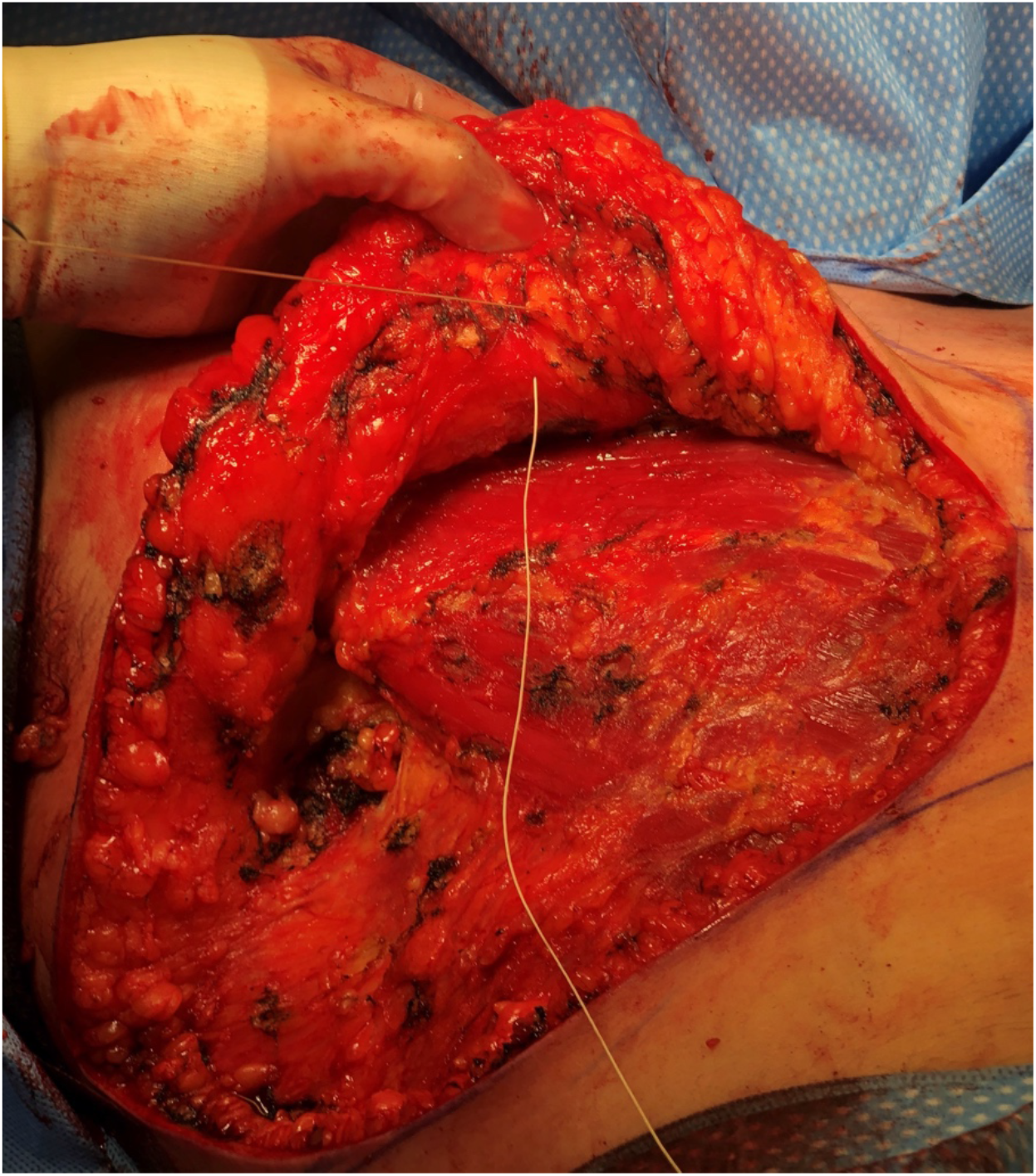
0-0 Vicryl suture placed through the superficial fascia, closing dead space in a simplified manner.

**Figure 3.**
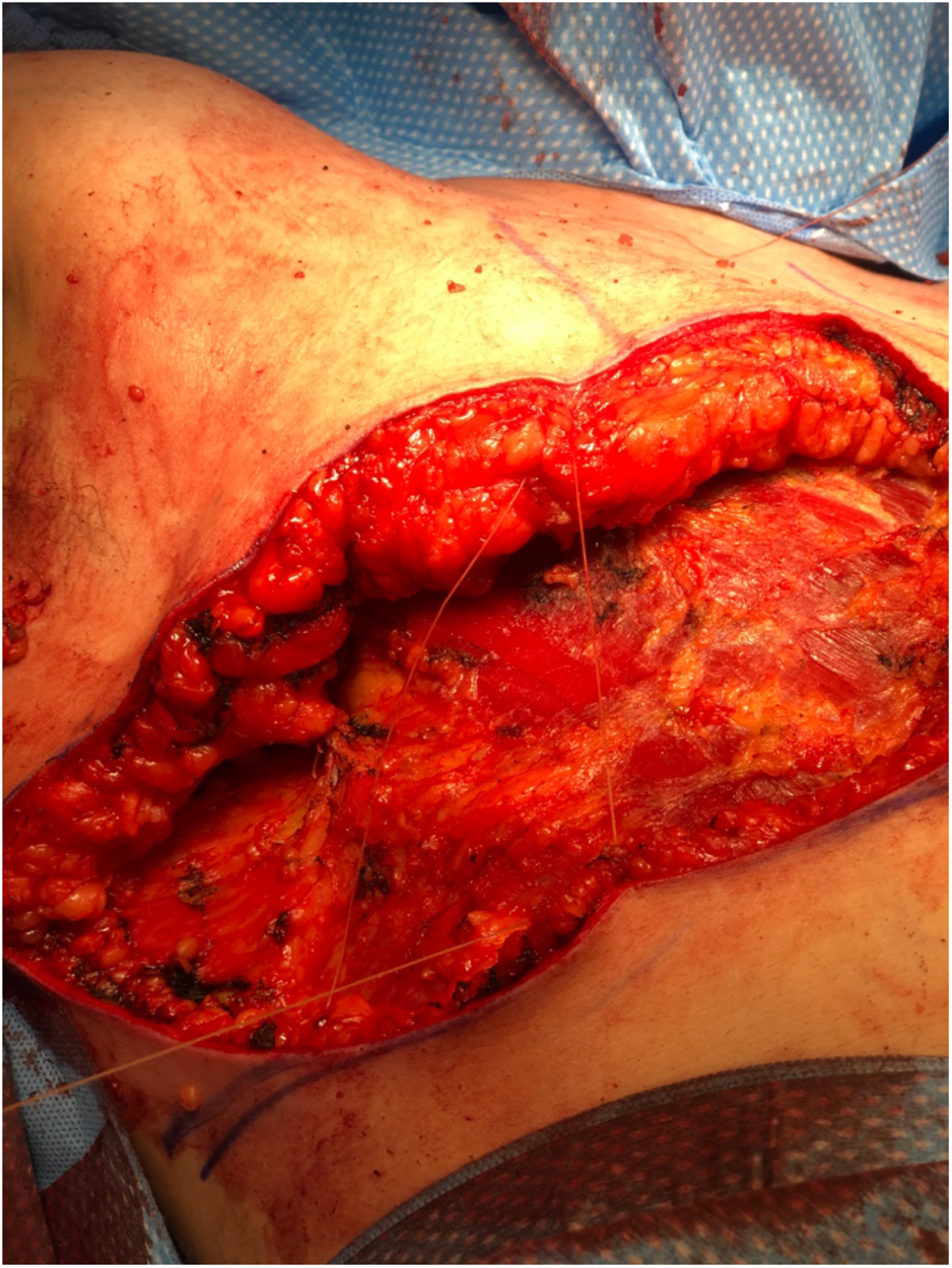
After closure of the superficial fascia with 4 0-0 Vicryl sutures.

**Figure 4.**
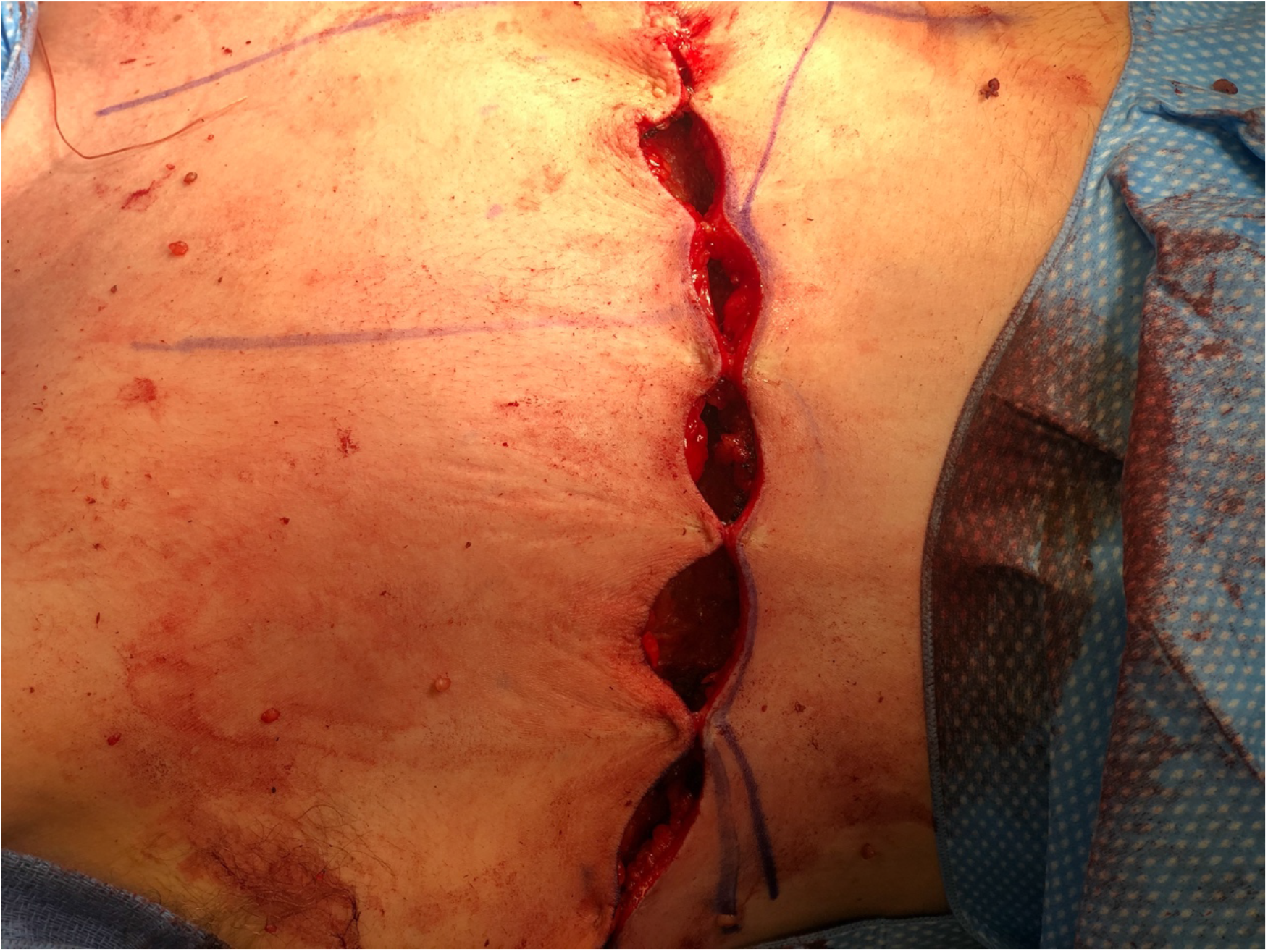
Closure of wound with the Insorb® stapler.

**Figure 5.**
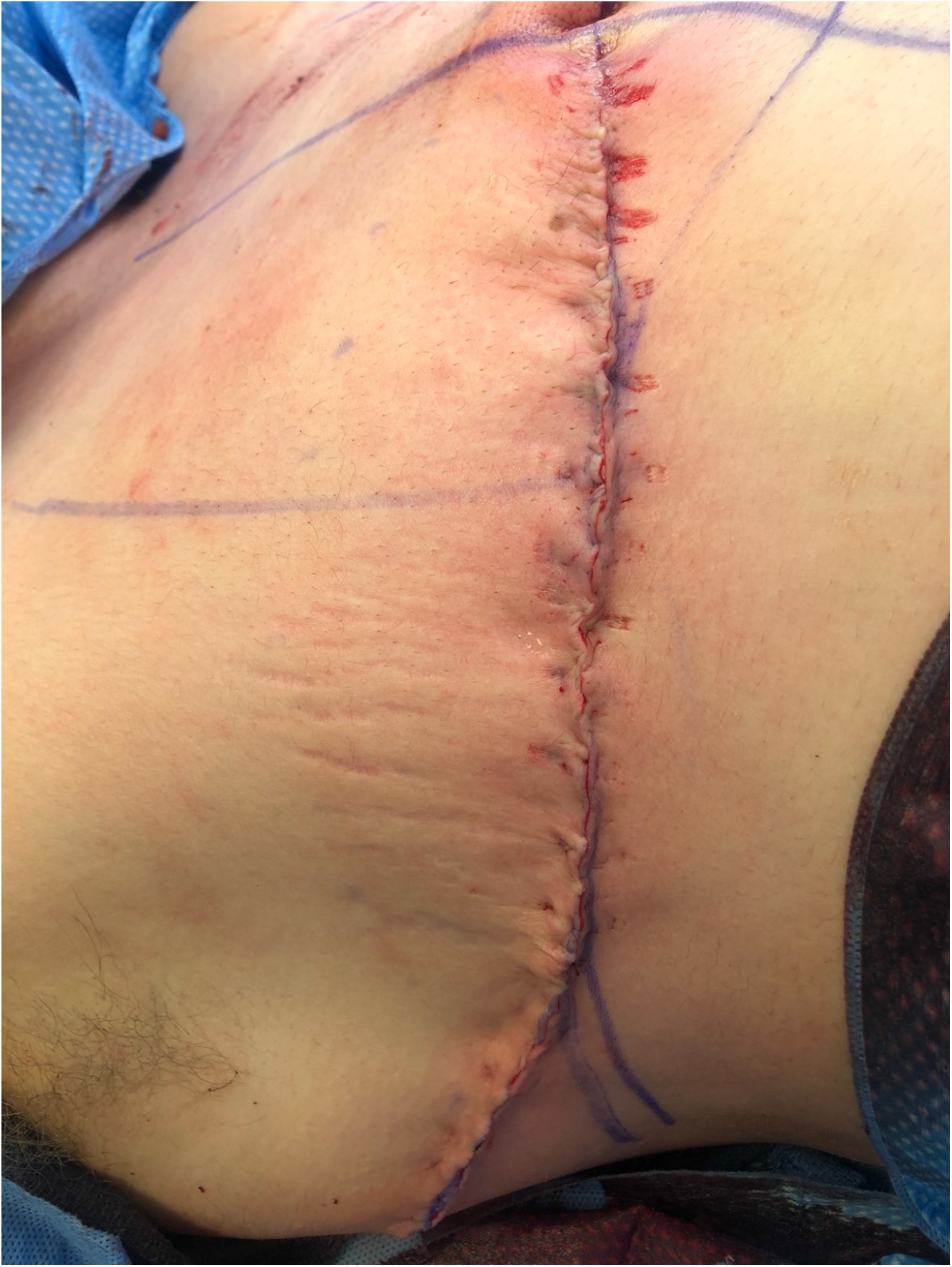
After complete closure.

All surgery is outpatient and patients are discharged from the Post-Anesthesia Care Unit to home after satisfactory recovery, usually in less than an hour. The abdominal binder and nipple bolster dressings are removed on postoperative day #5. Patients are instructed not to lift more than 20 pounds (9 kg) for 3 weeks after surgery. Patients are given the option to follow up a second time, but most elect to skip this appointment. Close follow-up via phone, text, or email is maintained with the patient.

## Results

### Patient Demographics

Between August 2017 and June 2020, 218 consecutive subcutaneous mastectomies were performed on 109 patients. Two patients were excluded from the study as they were treated using keyhole circumareolar incision. The remaining 107 patients (214 breasts) underwent a simplified DIFNG technique without drain placement. Mean patient age was 27.2 ± 10.4 years (13 – 60 years). 70% of the primary procedures were paid for by insurance. In these patients, the average co-pay was $12 ± $20 ($0 – $60).

Our patients were 65.4% White, 16.3% Hispanic, 7.2% Asian, 5.3% African American, 3.8% American Indian, and 1.9% Other. Upon consultation, 29.9% of our patients had a history of smoking and were encouraged to stop three months prior to surgery. By the date of surgery 51.4% of our patients had least 1 year of continuous testosterone therapy. Four patients had history of thyroid disease, two had history of diabetes, two had history of lung disease, one had history of heart disease, and one had history of cancer. Our patients had a mean BMI of 26.7 ± 6.3 kg/m^2^ (16.6 – 48.1 kg/m^2^) and 26.5% of our patients were obese.

### Intraoperative

After surgery, our patients had a mean estimated blood loss of 53 ± 29 cc (5 – 200 cc). From resulting pathology, mean right breast resection was 473 ± 291 g (92 – 1288 g), while the mean left breast resection was 471 ± 288 g (96 – 1332 g).

### Postoperative Outcomes

The overall complication rate was 13.1 percent, with a major complication rate of 0.9 percent, and a minor complication rate of 12.1 percent. The only major complication was a hematoma (1/107, 0.9%) that occurred early in our experience, which required return to the operating room. Minor complications affected 13/107 cases (12.1%) but were self-limited or treated in the clinic in all cases. Of the 10/107 (9.3%) cases of seroma, only 2/107 (1.9%) required aspiration, while the other eight resolved on their own with no further intervention. The remaining minor complications of one minor hematoma and two instances of wound dehiscence resolved on their own without further intervention by our surgeons. There was no loss to follow-up.

The overall elective revision rate was 3/107 (2.8%). The cause of revision was: scar reduction (1 patient), liposuction for asymmetry (1 patient), and lateral wound dog ear reduction (1 patient). The overall revision rate, including one acute reoperation for hematoma evacuation was 4/107 (3.7%).

The outcomes of our cohort were compared with previously published outcomes studies of FTM top surgery using Fisher’s exact test. Our literature review resulted in eleven studies with drain placement (Table 1) and two studies without drain placement (Table 2). When our drainless cohort was compared with pooled historical outcomes of patients with drain placement, data-analysis revealed that our drainless group had significantly higher rates of seroma (*p* = 0.003353), but significantly lower rates of revision (*p* = 1.37×10^−12^). Rates of hematoma, wound dehiscence, SSI, and NAC necrosis were found not to be significantly different (*p* = 0.07591; 1.000; 0.1367; 0.2377).

**Table 1:**
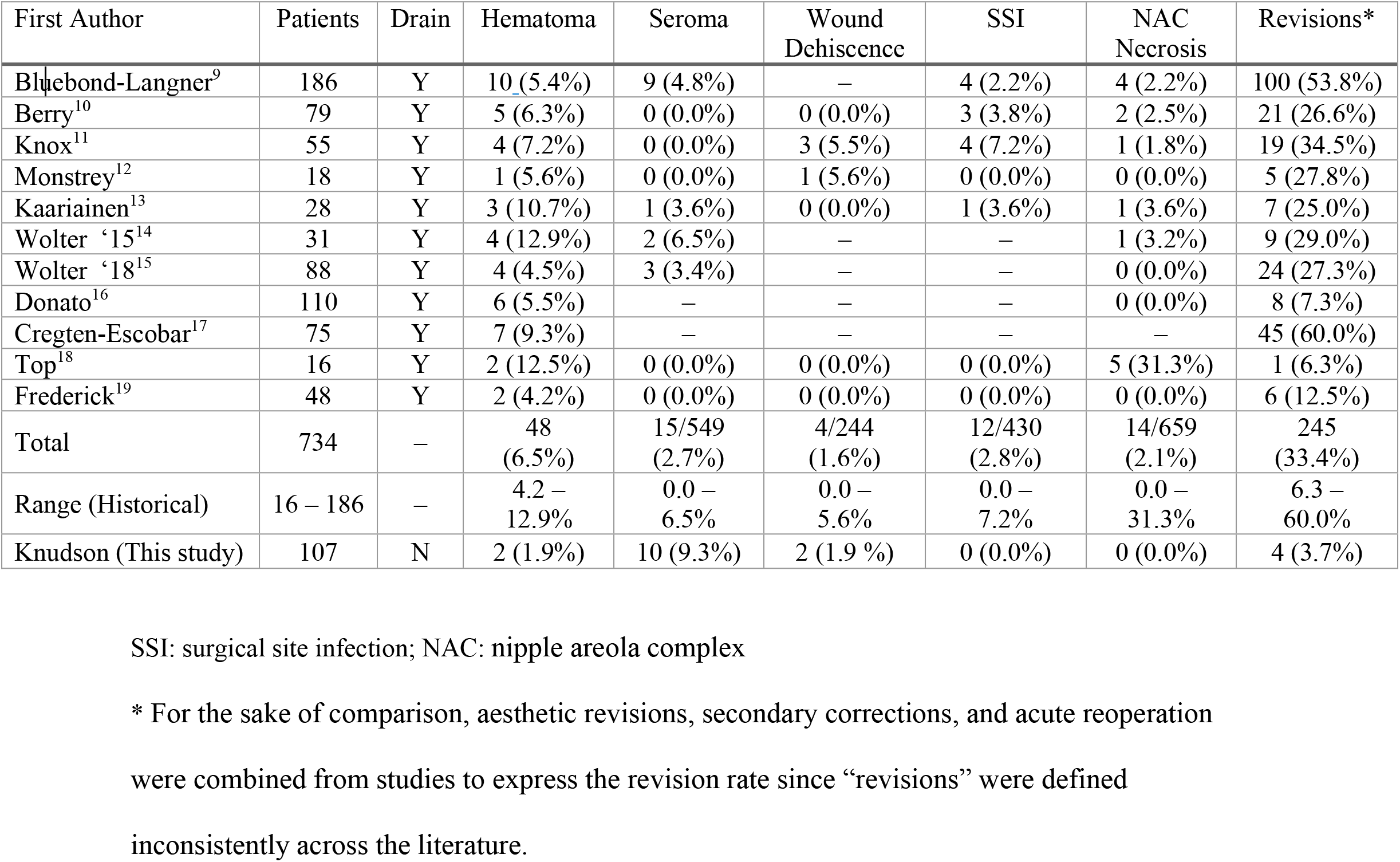
Historical drain inclusive outcome studies for DIFNG technique

**Table 2:** Historical drainless outcome studies for DIFNG technique

| First Author | Patients | Surgical Method | Hematoma | Seroma | Wound Dehiscence | SSI | NAC Necrosis | Revisions* |
| --- | --- | --- | --- | --- | --- | --- | --- | --- |
| Gallagher<sup>21</sup> | 153 | Obliteration | 1 (0.6%) | 0 (0.0%) | 3 (2.0%) | 7 (4.6%) | 2 (1.3%) | 9 (5.9%) |
| McEvenue<sup>22</sup> | 575 | Obliteration | 22 (3.8%) | 25 (4.3%) | 3 (0.5%) | 20 (3.5%) | 3 (0.5%) | 66 (11.5%) |
| Total | 728 | – | 23 (3.2%) | 25 (3.4%) | 6 (0.8%) | 27 (3.7%) | 5 (0.7%) | 75 (10.3%) |
| Range (Historical) | 153 – 575 | – | 0.6 – 3.8% | 0.0 – 4.3% | 0.5 – 2.0% | 3.5 – 4.6% | 0.5 – 1.3% | 5.9 – 11.5% |
| Knudson (This study) | 107 | Simplified | 2 (1.9%) | 10 (9.3%) | 2 (1.9 %) | 0 (0.0%) | 0 (0.0%) | 4 (3.7%) |

SSI – surgical site infection; NAC – nipple areola complex
\* For the sake of comparison, aesthetic revisions, secondary corrections, and acute reoperation were combined from studies to express the revision rate since “revisions” were defined inconsistently across the literature.

The three drainless studies, including our study, (Table 2) were then pooled together and compared with the eleven drain inclusive studies, data analysis showed slightly different results. Pooled drainless outcome data revealed significantly lower rates of hematoma (*p* = 0.001069), NAC necrosis (*p* = 0.01034), and revision (*p* = 2.20×10^−16^). Rates of seroma, wound dehiscence, and SSI were not significantly different (*p* = 0.1853; 0.4845; 0.7338).

Among the drainless studies, Gallagher et al. and McEvenue et al. utilized an obliterative surgical method while our study used simplified suturing. Gallagher et al. and McEvenue et al. were pooled together and were compared with our study. Data analysis revealed that our simplified method had significantly higher rates of seroma (*p* = 0.008911), but significantly lower rates of SSI (*p* = 0.03840) and revision (*p* = 0.03221). Rates of hematoma, wound dehiscence, and NAC necrosis were not significantly different (*p* = 0.7597; 0.2734; 1.000).

## Discussion

Drainless, outpatient, simplified DIFNG technique using a streamlined method of eliminating dead space can be performed with low rates of complication, especially complications that are generally addressed with drain placement: hematoma, seroma, dehiscence, SSI, NAC necrosis, and revisions. Our 1.9% rate of hematoma compares favorably to published rates of 4.2 **–** 12.9% when drains are used,^9,10,19,11–18^ and 0.6 **–** 3.8% when they are not used.^21,22^ Additionally, analysis revealed that drainless studies had significantly lower rates of hematoma than studies that used drains (*p* = 0.001069). Yet, this significance was not realized during single study comparison against aggregate drain inclusive historical outcomes, nor when compared against aggregate drainless historical outcomes. Although not significant in two analyses, our rate is historically similar. Given our analysis and historical accounts, drainless methods of DIFNG are confirmed to reduce hematoma rates.^21^ The type of drainless DIFNG surgical method has not shown to differ regarding hematoma rate,^21,22^ yet patients are still significantly more likely to develop hematoma post DIFNG with a drain than without one.

A similar pattern was revealed in our analysis of NAC necrosis. Our 0.0% rate of NAC necrosis was comparable and on the lower end of the historical rate range of 0.0 – 31.3% in series that used drains,^9–16,18,19^ and of the historical rate range of 0.5 – 1.3% in series that did not use drains.^21,22^ Significantly lower rates of NAC necrosis were found when pooled drainless studies were compared with pooled drain inclusive studies (*p* = 0.01034). It can be concluded that drainless DIFNG surgery reduces rates of NAC necrosis. Additionally, the type of drainless DIFNG surgery has not shown to significantly differ the rate of this complication.

Our 9.3% rate of seroma (1.9% requiring aspiration) was higher than generally reported rates of 0.0 – 6.5% in series that use drains,^9–15,18,19^ and 0.0 – 4.3% when drains were not used.^21,22^ Given that our rate of seroma was above expected ranges, data analysis revealed our rate was significantly higher than pooled drain inclusive historical outcomes (*p* = 0.003353) and pooled drainless outcomes (*p* = 0.008911). Although when pooled with drainless studies and compared against the aggregate drain inclusive data, no significance was realized. We can hypothesize that our streamlined obliteration of dead space may allow higher potential for seroma formation, although these seromas appear to only occasionally require treatment (one-time bedside aspiration and one aspiration conveniently during revision surgery). A majority of our reported seromas were minor and resolved on their own.

Our 1.9% rate of wound dehiscence was within the historical rate range of 0.0 – 5.6% in series that used drains,^10–13,18,19^ and rate range of 0.5 – 2.0% in series that didn’t use drains.^21,22^ Additionally there were no significant differences among dehiscence rates when comparing the aggregate historical data to our study. From these findings we can conclude that not using a drain makes little difference in resultant rate of wound dehiscence.

On the other hand, our 0.0% rate of SSI was found to be significantly lower than rates in studies that didn’t use drains (*p* = 0.0384) (3.5 – 4.6%),^21,22^ but not significantly different from rates in studies that used drains (0.0 – 7.2%).^9–13,18,19^ This result can be attributed to our cohort having zero instances of SSI.

Furthermore, our need for revision surgery (3.7%), is similar to that reported by surgeons using drains (6.3 – 60.0%),^9,10,19,11–18^ and those not using drains (5.9 **–** 11.5%).^21,22^ There is wide variation in this variable in the literature. This can be attributed to recommendations and preferences for revisions by specific surgeons. Although significance was found for this variable, it is unlikely that drain placement has much influence over the resultant rate.

Both major reports on the results of drainless DIFNG^21,22^ in the literature emphasize a technique of thorough obliteration of dead space using a method described for drainless abdominoplasty.^23,24^ It is described by Gallagher et al. that using 1-0 Vicryl sutures through the superficial fascia of the superior flap, then through caudal pectoralis fascia to leave “minimal dead space” throughout the surgical field.^21^ It appears to achieve equivalent results with only 4 deep flap sutures, 4 to 6 additional deep sutures at the skin incision, assisted and speeded by the use of deep and superficial absorbable staples.

### Minimal Impact Surgery

This surgery is successfully and safely performed on an outpatient basis, without drains, with only one follow-up appointment needed in most cases, and only 5 days of postoperative binder use. This sort of “minimal impact” surgery system has been advocated previously to streamline surgery, improve patient experience, decrease pain, lower costs, and lower some patient barriers to completing surgery. Similar outpatient minimal impact regimes (including eschewing drains in the case of male urethroplasty) have proven successful in: complicated urethral surgery,^25^ major joint arthroplasty,^26^ umbilical hernia repair,^27^ and other major surgery types. We and others^19,21^ have shortened postoperative use of compression garments significantly, to 5-7 days, which is a significant reduction from older reports that require 3-6 weeks of compression.^10,11,13,15,16^ While no data exists to quantify the exact patient benefit of 2-5 weeks less compression garment use, it is famously uncomfortable, inconvenient, and detested among our patients. Allowing the patient to be free of extended chest compression after surgery has been received by our patients with much satisfaction.

### Benefits of Eliminating Drains

The many negative aspects of drains are well-reviewed,^21^ but can be briefly summarized here: avoidance of scar at exit site, subjective reports of easier post-operative care regimen for patients,^23^ less surgical pain,^28^ and even decreased hospital length of stay in certain situations.^29^ As with binders, no data exists to exactly quantify the benefits of avoiding drains, but allowing the patient to avoid the bother and discomfort of drains will be perceived by them as a major benefit.^23^ Additionally drains are sites of bacterial colonization, as SSI post bilateral mastectomy has been shown to decrease with drain antisepsis.^30^ In our analysis, SSI was found not significantly different than historical outcomes, further indicating drains are not needed for low rates of complication following DIFNG.

### Limitations

Limitations of this study include our retrospective approach, lack of patient reported outcomes, and short follow-up. Additionally, data analysis was limited due to different sample sizes of pooled groups during comparison.

## Conclusions

With the expansion of coverage and societal acceptance of transgender surgical services, FTM transitions have become more prevalent in the United States population. Therefore, further outcome literature is needed to distinguish best practice. We report that simplified, drainless, outpatient, DIFNG mastectomy can be safely offered. We reported a comparable complication rate, yet our simplified technique resulted in increased rates of seroma relative to past drainless studies. Although a majority of our patients’ seromas resolved on their own, this trend should be noted. In comparison to historical data, we confirm that abandoning the practice of drain placement decreases rates of hematoma, NAC necrosis, and revision.

## Data Availability

All data produced in the present study are available upon reasonable request to the authors

